# Association Between Hospital Tiers and Cardiogenic Shock Mortality: Mitigating the Transfer Penalty Through a Regionalized Hub-and-Spoke Model

**DOI:** 10.64898/2026.04.05.26350211

**Authors:** Ankur Sethi, Emily Hiltner, Ashish Awasthi, Casey Panebianco, Tana LaPlaca, Nancy Rizzuto, Leonard Y Lee, Mark Russo

**Author notes:** Corresponding author – Ankur Sethi MBBS FACC FSCAI, 901 West Main Street, Freehold, NJ 07728, Email-, Twitter- @AnkurSethi2021.

## Abstract

**Background:** Cardiogenic shock (CS) remains associated with high short-term mortality despite contemporary advances in care. The association between institutional cardiac capability and outcomes—particularly among transferred patients and after accounting for clinical instability—remains incompletely defined.

**Objectives:** To evaluate the association between hierarchical hospital cardiac capability and in-hospital mortality using a latent measure of acute physiologic severity.

**Methods:** Using the National Inpatient Sample (2016–2022), hospitals were classified into five hierarchical tiers ranging from non-PCI (Tier 1) to heart transplant/durable LVAD centers (Tier 5). Generalized structural equation modeling (GSEM) assessed the relationship between hospital tier and mortality. A latent “Acute Severity” construct—comprising cardiac arrest, acute kidney and liver injury, and mechanical ventilation—was incorporated to model the effects of clinical instability

**Results:** Among an estimated 1,177,180 CS hospitalizations, most occurred at cardiac surgical and transplant/LVAD centers. Crude mortality declined stepwise from non-PCI hospitals (64.5%) to transplant/LVAD centers (36.5%). After adjustment, higher hospital tier was independently associated with lower mortality (Tier 2 OR 0.43 [95% CI 0.38–0.48]; Tier 3 OR 0.37 [0.32– 0.43]; Tier 4 OR 0.33 [0.30–0.38]; Tier 5 OR 0.35 [0.31–0.40]). Although transfer-in status was associated with increased mortality (OR 1.39 [1.33–1.46]), this association was attenuated at cardiac surgical and transplant/LVAD centers, consistent with a mitigation of transfer associated risk.

**Conclusions:** Higher hospital cardiac capability is independently associated with lower mortality in CS. Advanced centers are associated with mitigation transfer-associated risk, supporting regionalized hub-and-spoke systems with early referral to high-capability centers.

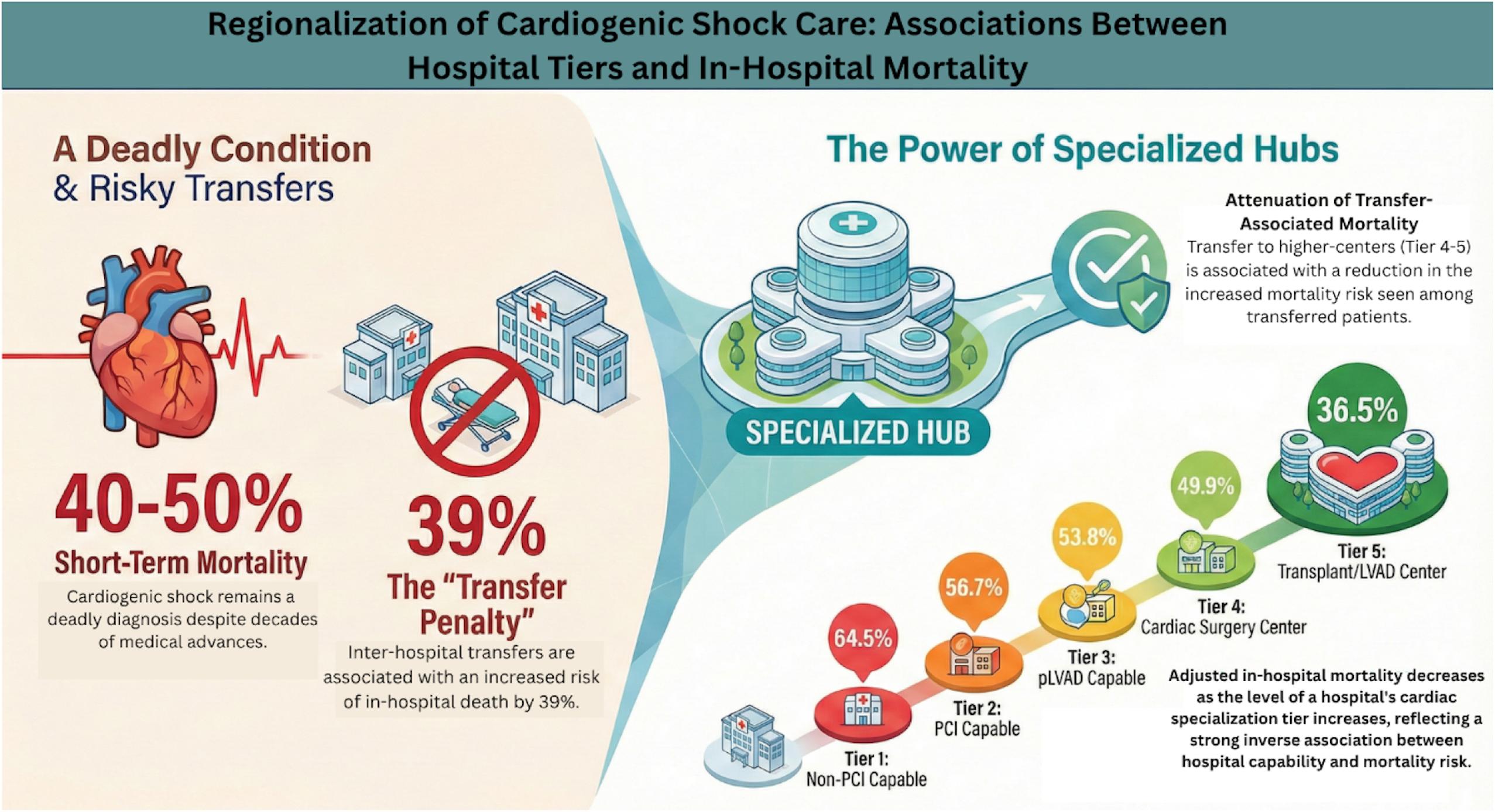

## Background

Cardiogenic shock (CS) is a complex clinical syndrome defined by end-organ hypoperfusion secondary to inadequate cardiac output. Despite decades of cardiovascular advancement, short-term mortality remains stubbornly high at 40–50%, often characterized by significant regional and institutional variation^1 2 3^. To date, randomized controlled trials have identified only two interventions that definitively reduce mortality in ST elevation myocardial infarction (STEMI) related CS: early revascularization and, more recently, the use of micro-axial left ventricular flow pumps ^4 5^.

However, a significant gap remains between clinical evidence and practice. Widespread disparities exist across the United States in the utilization of primary percutaneous coronary intervention (PCI) and mechanical circulatory support (MCS)^6 7^. This challenge is further compounded by the evolving epidemiology of the disease; nearly two-thirds of contemporary CS cases are now non-acute myocardial infarction(AMI) related^8^. Many primary PCI centers lack the specialized cardiac intensive care, multidisciplinary heart teams, and advanced MCS capabilities required to manage these increasingly complex patients. Given the time-sensitive nature of the condition, the American Heart Association has proposed a regionalized ‘hub-and-spoke’ system of care to optimize the triage and transfer of CS patients to specialized tertiary centers“^9^. Although a regionalized approach is conceptually logical—aligning with established data on the volume-outcome relationship in complex cardiac care—the specific impact of institutional cardiac capabilities on CS mortality has not been rigorously evaluated at a national level. Furthermore, a critical paradox exists; while specialized centers offer superior resources, the process of inter-hospital transfer has been independently associated with increased mortality in CS patients^10^. Understanding the intricate interaction between a hospital’s specialized capabilities and the inherent risk of transfer is vital before the United States moves toward the widespread adoption of a hub-and-spoke model. Importantly, prior national analyses using administrative data have been unable to account for unmeasured clinical instability, a limitation that may obscure true institutional effects. This study seeks to bridge that gap by leveraging a latent variable modeling framework to quantify acute physiologic severity, allowing a more accurate comparison of outcomes across hospital tiers while adjusting for the high acuity typical of transferred patients.

## Methods

### Data Source and Study Population

Data were derived from the National Inpatient Sample (NIS) for the years 2016 through 2022. Sponsored by the Agency for Healthcare Research and Quality (AHRQ) as part of the Healthcare Cost and Utilization Project (HCUP), the NIS is the largest all-payer inpatient database in the United States. Since 2012, the NIS has employed a stratified sampling design that approximates a 20% systematic sample of all discharges from U.S. community hospitals, representing more than 97% of the national population.

We included all hospitalizations for adults (age ≥ 18 years) with a primary or secondary diagnosis of cardiogenic shock (CS), identified using the International Classification of Diseases, Tenth Revision (ICD-10) code R57.0. We excluded visits with missing data on vital status at discharge.

### Hospital Capability Classification

Hospitals were classified annually based on procedural capabilities. A hospital was considered capable of a given intervention if it performed that procedure at least once during the calendar year. Using this approach, hospitals were categorized into hierarchical tiers reflecting increasing levels of cardiovascular capability:

1. Non–PCI-capable hospitals
2. PCI-capable hospitals
3. Percutaneous left ventricular assist device (pLVAD)–capable hospitals
4. Cardiac surgery (Coronary artery bypass-capable) centers
5. Heart transplant and/or durable LVAD–capable centers

This tiered framework reflects real-world escalation pathways for patients with cardiogenic shock.

### Covariates and Clinical Characteristics

Patient demographics and comorbidities were identified using ICD-10 diagnosis codes. Comorbid conditions were defined using the Elixhauser comorbidity index based on the coding algorithm developed by Quan et al.^11^, including hypertension (with and without complications), diabetes mellitus (with and without complications), congestive heart failure, cardiac arrhythmias, and valvular heart disease. Overall comorbidity burden was quantified using the weighted Elixhauser comorbidity summary score (Elixsum).

We additionally identified hospitalizations complicated by cardiac arrest, AMI, acute kidney injury (AKI), acute liver injury (ALI), and use of invasive mechanical ventilation. Utilization of cardiovascular procedures and support devices—including right heart catheterization, intra-aortic balloon pump, pLVAD, and extracorporeal membrane oxygenation (ECMO)—was determined using ICD-10 procedure codes (Table S1 in the supplement).

### Outcomes

The primary endpoint was in-hospital mortality. Because discharge vital status is unknown for hospitalizations transferred to another acute care facility, these visits were excluded from mortality analyses.

## Statistical analysis

The analysis was performed using survey (svy) commands with appropriate sampling weights provided by the NIS, accounting for the stratification and clustering to obtain national estimates. The categorical variables were presented as proportions (percentages) and continuous variables were presented as means with their respective 95% confidence intervals or standard errors.

We employed a Generalized Structural Equation Model (GSEM) with a Bernoulli distribution and logit link function to examine the relationship between hospital capability tiers and in-hospital mortality. This framework was chosen over traditional multivariable logistic regression for three primary reasons: its ability to model unobserved (latent) patient states and quantify the mechanisms (paths) through which hospital infrastructure impacts survival (Figure 1). To address the multidimensional nature of patient instability and minimize multicollinearity among highly correlated clinical complications, we constructed a latent variable termed “Acute Severity” constructed from well-known complications of cardiogenic shock namely, ALI, AKI, use of invasive ventilation, occurrence of cardiac arrest. This accounts for the ‘delayed presentation’ effect, where transferred patients may arrive in a more advanced state of physiological collapse. The GSEM was structured to partition the Total Effect of hospital capability into two distinct pathways. The Direct effect attributable to institutional expertise and specialized care independent of the patient’s measured complications and Indirect effect achieved through the hospital’s ability to stabilize or mitigate the Acute Severity latent variable. To ensure nationally representative estimates, the model was nested within a survey estimation framework (svy) using National Inpatient Sample (NIS) probability weights. Variance was estimated using Taylor-series linearization to account for the complex sampling design, including stratification and clustering within hospitals. All models were adjusted for age, sex, and chronic illness burden via the standardized Elixhauser Comorbidity Index. We also adjusted for hospital-level confounders, including bed size and teaching status, to isolate the effect of cardiac-specific capabilities from general institutional characteristics. Furthermore, we investigated the interaction between hospital tier and transfer status using a formal interaction term to determine if higher-capability centers mitigated the mortality risk associated with inter-hospital transfer.

**Figure 1.**
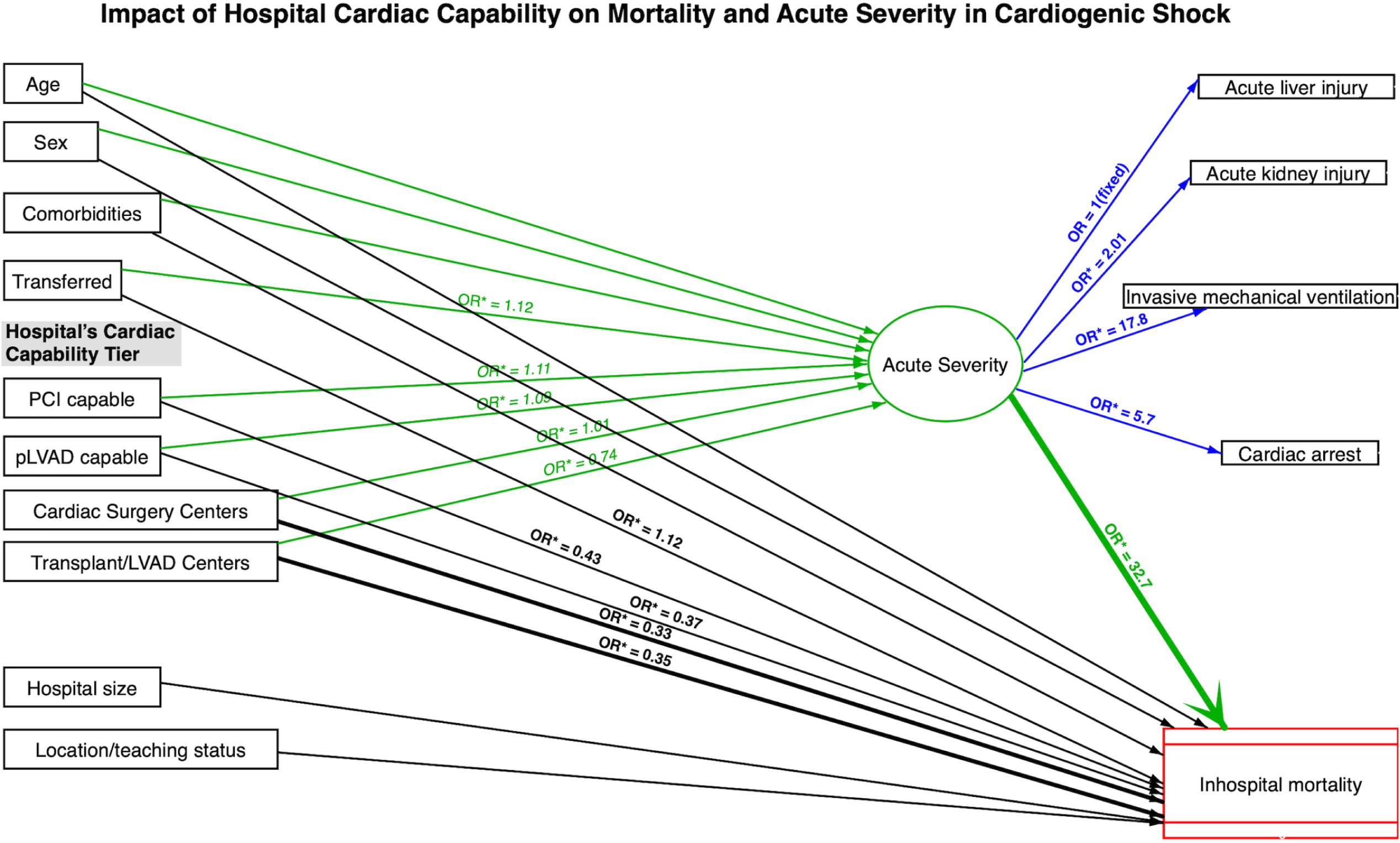
Structural Equation Model of Hospital Capability and Cardiogenic Shock Mortality. Path diagram illustrating the relationship between institutional capability, latent patient severity, and in-hospital survival. Ovals represent the latent construct of “Acute Severity“; rectangles represent observed variables. Green paths indicate factors contributing to acute physiological collapse (e.g., transfers and comorbidities). Thickened black paths highlight the primary direct effects of hospital tiers. Values on paths to mortality represent Odds Ratios (OR); values on measurement paths represent exponentiated factor loadings. (* p<0.05).

A subgroup analysis was performed for visits with AMI related and non-AMI related CS. Model discrimination was assessed using the Area Under the Receiver Operating Characteristic (AUROC) curve, derived from the predicted probabilities of the survey-weighted GSEM. To ensure the stability of our findings, a series of sensitivity analyses were performed. First, we compared GSEM results to standard survey-weighted multivariable logistic regression to confirm the directionality and magnitude of the primary associations. Second, to address potential misclassification and ensure hospital tiers reflected established systems of care rather than sporadic procedural capability, we executed an additional sensitivity model using stringent annual volume-based thresholds. In this model, hospital classification required a minimum of ≥ 10 PCI procedures, ≥ 5 CABG procedures, ≥ 5 pLVAD procedures, and ≥ 2 durable LVAD or heart transplant procedures during the calendar year.

This study was exempt from institutional review board approval because the NIS is a publicly available, de-identified administrative database. All analyses adhered to AHRQ-recommended methodological standards for research using the NIS^12^.

## Results

### Baseline Patient Characteristics and Comorbidities

A total estimated 1,177,180 visits for CS were analyzed across five hospital capability tiers. The largest proportion of patients were treated at Tier 4 centers (44.9%), followed by Tier 5 centers (34.5%). Patients treated at Tier 5 centers were significantly younger (mean age 63.5 years) than those at Tier 1 centers (mean age 70.0 years; p<0.0001) and were more frequently male.

Overall comorbidity burden was high across all tiers but varied significantly by institutional capability. Tier 5 centers managed patients with the highest prevalence of congestive heart failure (81.3%) and valvular heart disease (31.3%), resulting in the highest mean Elixhauser comorbidity index (6.51; p<0.0001) (Table 1).

**Table 1.** Characteristics of cardiogenic shock visits to different hospital tiers. Tier 1 – non PCI capable, Tier 2 – PCI capable, Tier 3 – pLVAD capable, Tier 4 – Cardiac surgery centers, Tier 5 – Transplant/LVAD centers.

| *Characteristic* | *Tier 1* | *Tier 2* | *Tier 3* | *Tier 4* | *Tier 5* | *P value* |
| --- | --- | --- | --- | --- | --- | --- |
| **Total Estimated Cases (N)** | 58,575 | 127,430 | 57,005 | 528,040 | 406,130 | N/A |
| **Population Proportion (%)** | 4.98 | 10.82 | 4.84 | 44.86 | 34.5 | N/A |
| **Age, Mean (SE)** | 70.02 (.15) | 68.25 (.11) | 67.96 (.18) | 67.45 (.06) | 63.48 (.10) | <0.001 |
| **Categorical Variables, %** |  |  |  |  |  |  |
| Female | 43.56 | 40.62 | 38.91 | 37.64 | 35.1 | <0.001 |
| Transferred from another healthcare facility | 10.69 | 10.07 | 10.46 | 21.16 | 39.39 | <0.001 |
| **Race** |  |  |  |  |  | <0.001 |
| White | 65.57 | 66.84 | 67.55 | 70.1 | 63.34 |  |
| Non-White | 34.43 | 33.16 | 32.45 | 29.9 | 36.66 |  |
| **Primary Payer** |  |  |  |  |  | <0.001 |
| Medicare | 67.52 | 63.59 | 63.46 | 62.56 | 55.54 |  |
| Medicaid | 13.87 | 12.68 | 11.98 | 11.98 | 13.53 |  |
| Private | 13.39 | 17.15 | 17.05 | 18.33 | 24.84 |  |
| Self-pay/Other | 5.21 | 6.58 | 7.51 | 7.13 | 0.61 |  |
| **Comorbidities (Present) (%)** |  |  |  |  |  |  |
| Congestive Heart Failure | 73.85 | 72.47 | 72.06 | 74.33 | 81.33 | <0.001 |
| Cardiac Arrhythmias | 63.39 | 65.09 | 66.26 | 67.34 | 68.4 | <0.001 |
| Valvular Disease | 20.96 | 20.55 | 19.87 | 25.54 | 31.27 | <0.001 |
| Diabetes, Uncomplicated | 12.04 | 11.9 | 11.63 | 10.94 | 8.65 | <0.001 |
| Diabetes, Complicated | 29.6 | 30.6 | 30.57 | 31.71 | 29.89 | <0.001 |
| Hypertension, Uncomplicated | 14.2 | 16 | 15.7 | 16.16 | 12.13 | <0.001 |
| Hypertension, Complicated | 60.5 | 59.37 | 60.49 | 61 | 62.03 | <0.001 |
| **Elixhauser Comorbidity Score, Mean (SE)** | 6.28(.02) | 6.15(.02) | 6.06(0.04) | 6.23(0.01) | 6.51(0.03) | <0.001 |
| **ACUITY AND PRESENTATION** |  |  |  |  |  |  |
| STEMI | 4.32 | 17.63 | 20.72 | 19.28 | 11.93 | <0.001 |
| NSTEMI | 19.09 | 18.92 | 19.22 | 20.55 | 14.07 | <0.001 |
| Cardiac Arrest | 15.43 | 22.31 | 24.22 | 22.16 | 15.1 | <0.001 |
| Acute Liver Injury | 17.08 | 19.12 | 19.65 | 19.31 | 21.44 | <0.001 |
| Acute Kidney Injury | 65.04 | 63.49 | 62.1 | 62.35 | 65.4 | <0.001 |
| Invasive Mechanical Ventilation | 44.88 | 49.36 | 48.3 | 48.39 | 39.01 | <0.001 |
| **HIGH-ACUITY INTERVENTION** |  |  |  |  |  |  |
| Right Heart Catheterization | 0.77 | 3.96 | 6.52 | 6.93 | 24.15 | <0.001 |
| Intra-aortic Balloon Pump | 0.05 | 9.17 | 8.56 | 12.52 | 14.5 | <0.001 |
| Percutaneous left ventricle assist device | 0 | 0 | 8.85 | 8.28 | 6.62 | <0.001 |
| Extracorporeal Membrane Oxygenation | 0.03 | 0.08 | 0.31 | 1.3 | 6.42 | <0.001 |
| **Hospital size** |  |  |  |  |  | <0.001 |
| Small | 47 | 33 | 29 | 12 | 1 |  |
| Medium | 33 | 40 | 33 | 30 | 9 |  |
| Large | 20 | 27 | 38 | 58 | 90 |  |
| **Hospital location** |  |  |  |  |  | <0.001 |
| Rural | 22 | 11 | 8 | 3 | 0 |  |
| Urban nonteaching | 29 | 31 | 31 | 17 | 0.9 |  |
| Urban teaching | 49 | 58 | 61 | 80 | 99.1 |  |

The prevalence of STEMI demonstrated a non-linear distribution, peaking at Tier 3 (20.7%) and Tier 4 (19.3%) centers, while Tier 5 centers reported the lowest STEMI prevalence among PCI-capable hospitals (11.9%). Non–ST-elevation myocardial infarction (NSTEMI) was consistently prevalent across Tiers 1–4 (18.9%–20.6%) but was notably lower at Tier 5 centers (14.1%).

### Acute Organ Dysfunction and Severity Indicators

Markers of acute organ dysfunction, which informed the latent construct of Acute Severity, increased with hospital capability tier. The prevalence of ALI was highest at Tier 5 (21.4%) and Tier 4 (19.3%) centers. AKI was highly prevalent across all tiers, peaking at 65.4% in Tier 5 centers.

Utilization of invasive mechanical ventilation was highest at Tier 2 (49.4%) and Tier 4 (48.4%) centers, whereas Tier 5 centers had the lowest utilization rate (39.0%). In contrast, cardiac arrest was most frequently documented at Tier 3 (24.2%) and Tier 4 (22.2%) centers (p<0.0002), with a lower prevalence observed at Tier 5 centers (15.1%).

### High-Acuity Interventions and Hospital Characteristics

Use of advanced hemodynamic monitoring and mechanical circulatory support increased sharply with institutional capability. Right heart catheterization was performed in 24.2% of Tier 5 hospitalizations compared with only 0.8% at Tier 1 centers. ECMO use was nearly absent in Tiers 1–3 but reached 6.4% in Tier 5 centers (p<0.0001).

Hospital characteristics differed substantially across tiers. Tier 5 centers were predominantly large (90%), urban teaching hospitals (99.1%), whereas Tier 1 centers were more frequently small (47%) or rural (22%) institutions (p<0.0001). Additional visit characteristics are summarized in Table 1.

### Unadjusted Mortality and Latent Severity Modeling

Unadjusted in-hospital mortality demonstrated a strong inverse relationship with hospital cardiac capability (Figure 2 and central illustration). Mortality was highest at Tier 1 centers (64.4%) and declined stepwise through Tier 2 (56.7%), Tier 3 (53.8%), and Tier 4 (49.9%), with the lowest crude mortality observed at Tier 5 centers (36.5%; P for trend <0.001).

**Figure 2.**
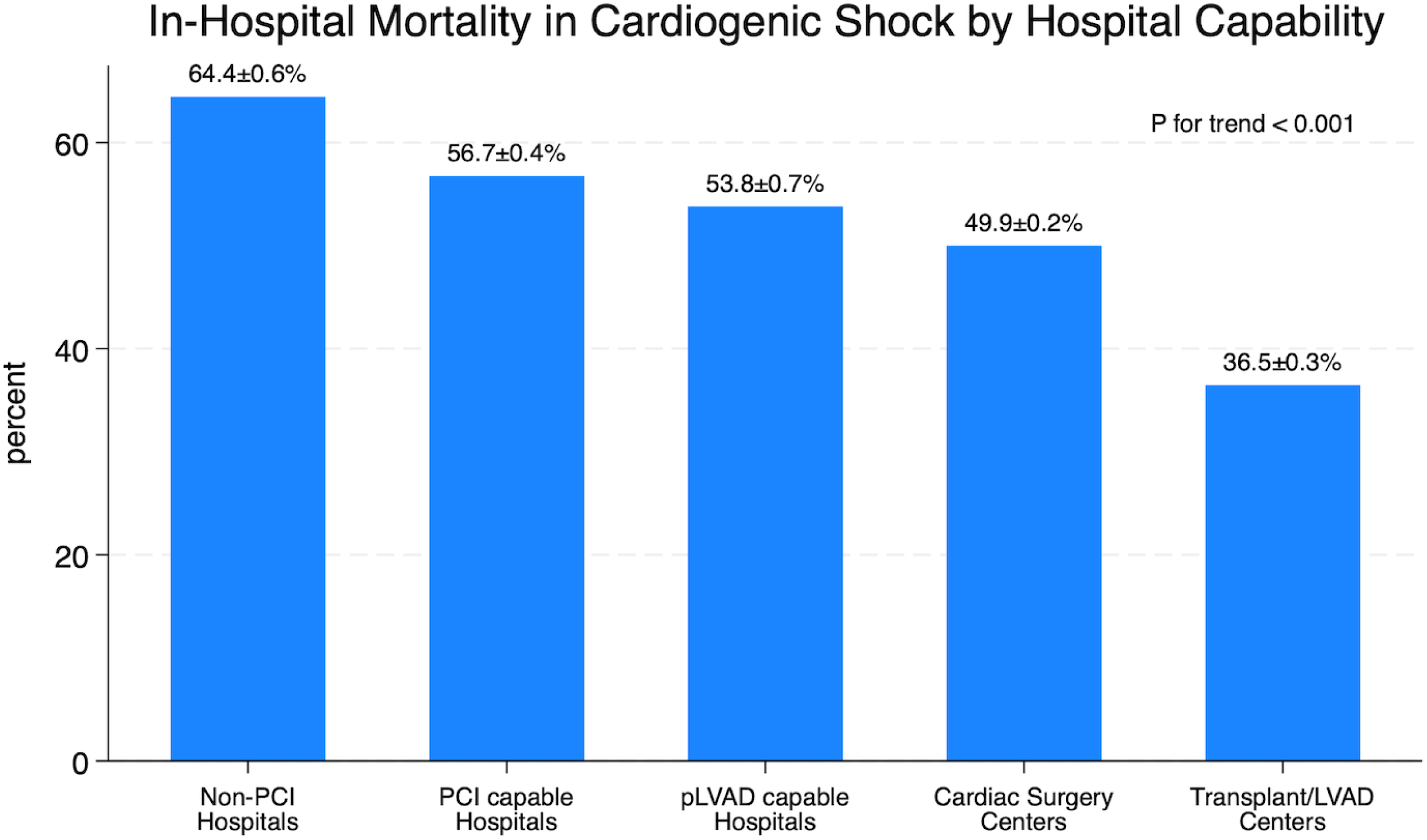
Unadjusted Mortality by Hospital Cardiac Capability. In-hospital cardiogenic shock mortality rates stratified by hierarchical hospital tiers. A significant stepwise reduction in crude mortality is observed as institutional capability increases. Error bars represent 95% confidence intervals.

However, direct comparison of crude mortality is limited by substantial selection bias, as Tier 5 centers treated a markedly higher-acuity population, characterized by a four-fold higher transfer-in rate (39.4%) and the inability of administrative data (NIS) to directly capture shock severity. To address this limitation, we employed a GSEM to define a latent construct of Acute Severity (Figure 1), serving as a mathematical proxy for underlying clinical instability.

This latent construct was strongly defined by AKI (OR 2.01; 95% CI 1.97–2.07), cardiac arrest (OR 5.70; 95% CI 5.22–6.21), and invasive mechanical ventilation (OR 17.8; 95% CI 15.27– 20.65). Transfer-in status (OR 1.12; 95% CI 1.10–1.14), greater comorbidity burden (OR 1.15; 95% CI 1.14–1.17), and younger age (OR 0.92; 95% CI 0.91–0.93) were significant upstream drivers of increased latent severity. Acute Severity emerged as the single strongest predictor of in-hospital mortality within the model (OR 32.7; 95% CI 28.83–37.21) as shown in Figure 1 and Table 2.

**Table 2.**
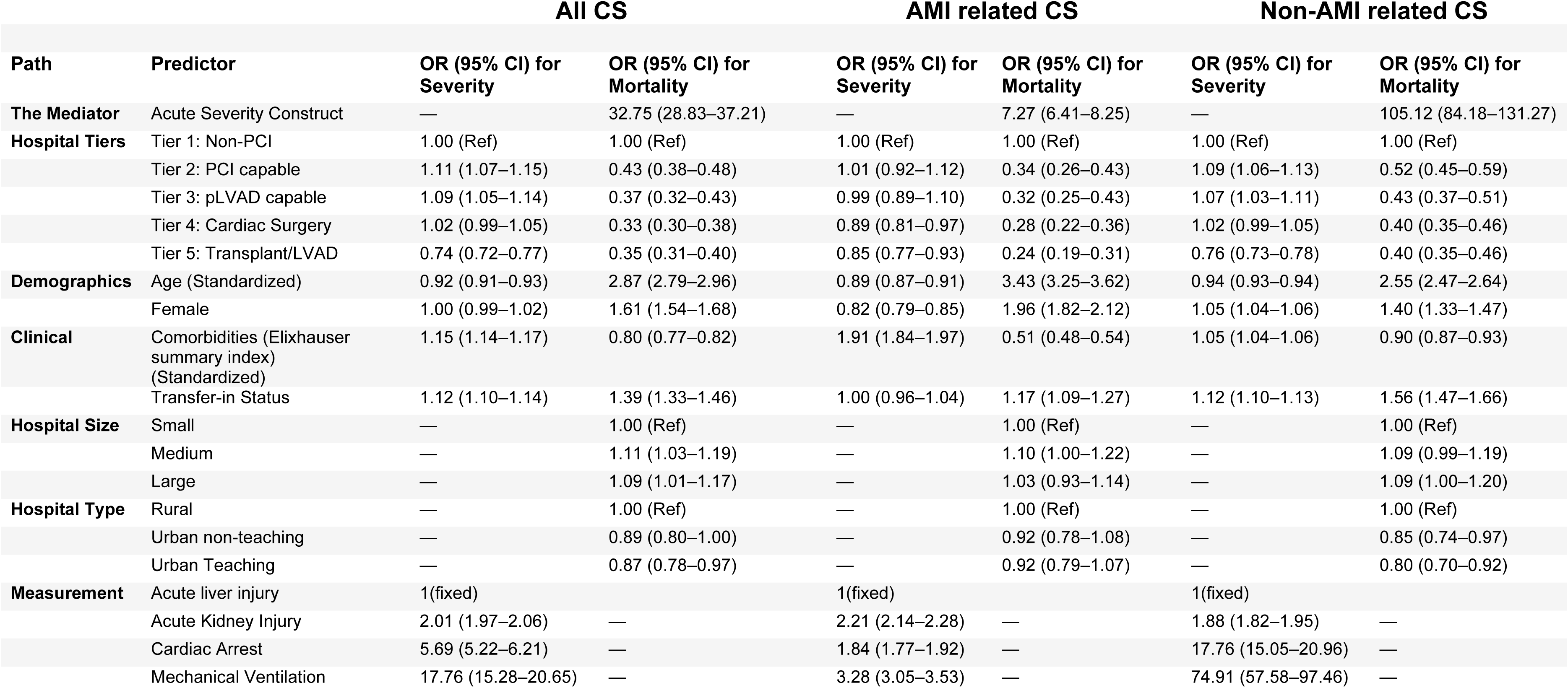
Survey-Weighted GSEM Path Coefficients for Cardiogenic Shock Outcomes. Detailed path coefficients for the total cohort, AMI-related, and non-AMI-related cardiogenic shock visits. Path for Severity columns represent odds ratios for the “Acute Severity” latent mediator, while Path for Mortality columns represent direct odds ratios for in-hospital mortality.

After adjustment for latent severity, age, sex, comorbidities, and hospital size and location, the survival advantage of higher-capability centers remained robust. Compared with Tier 1 hospitals, adjusted odds of in-hospital mortality were significantly lower at Tier 2 (PCI-capable; OR 0.43; 95% CI 0.38–0.48), Tier 3 (pLVAD; OR 0.37; 95% CI 0.32–0.42), Tier 4 (Cardiac surgery; OR 0.33; 95% CI 0.30–0.38), and Tier 5 (Transplant/LVAD; OR 0.35; 95% CI 0.31–0.40) centers (Figure 3 and Table 2).

**Figure 3.**
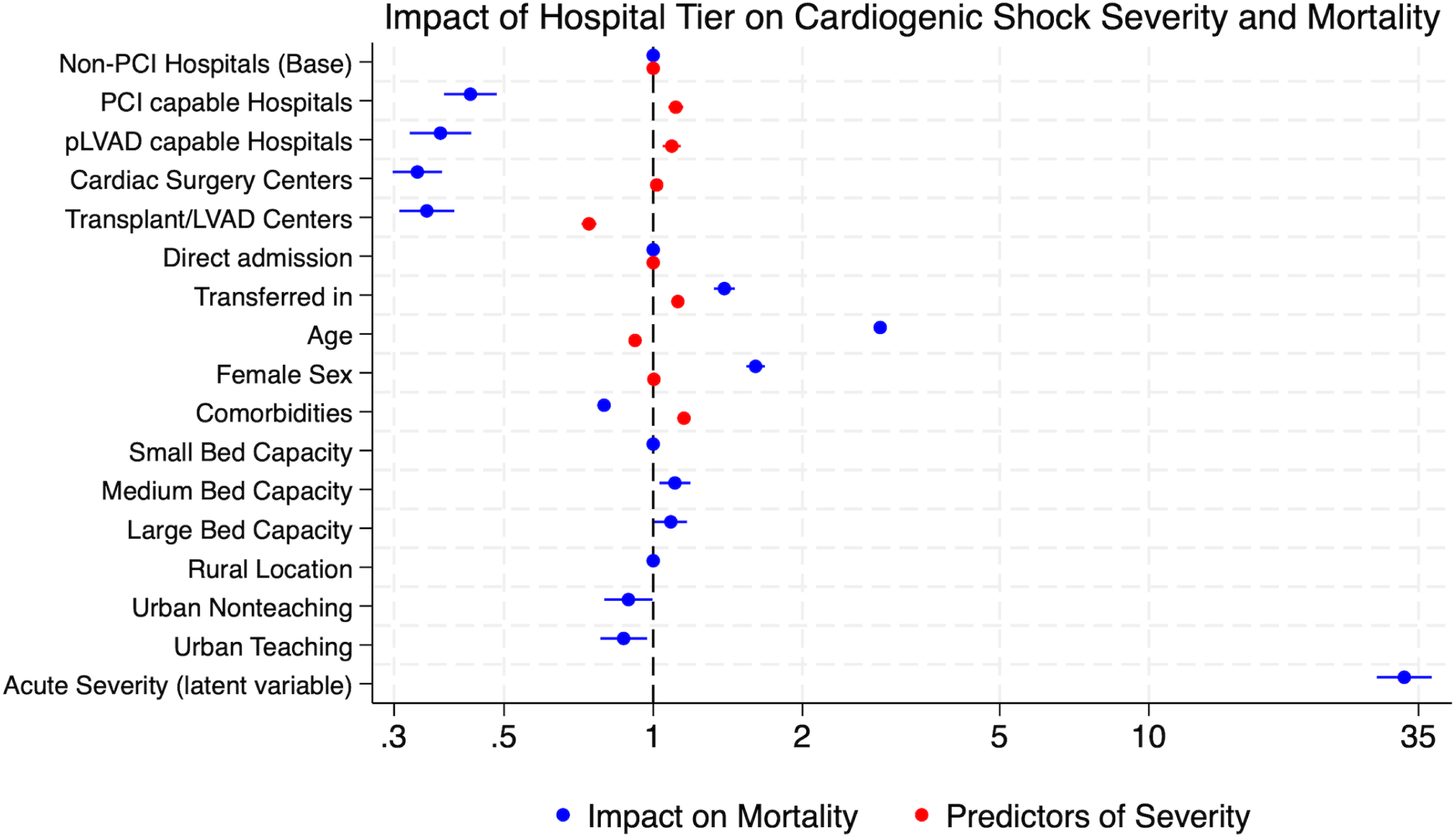
Institutional Capability, Latent Severity, and Mortality. GSEM results illustrating the drivers of shock mortality. Red dots represent predictors of the Acute Severity latent construct, while blue dots represent independent predictors of in-hospital mortality. Factors are categorized by hospital tier, patient characteristics, and institutional attributes. Error bars indicate 95% confidence intervals. LVAD - Left ventricle assist device(durable); PCI - Percutaneous coronary intervention; pLVAD - Percutaneous left ventricle assist device.

### Subgroup Analyses

#### AMI-Related Cardiogenic Shock

A total of 396,725 hospitalizations were for AMI-related CS, representing 33.7% (±0.2%) of all CS admissions. Unadjusted in-hospital mortality declined progressively from 75.5% at Tier 1 hospitals to 45.6% at Tier 5 centers (Figure 4A).

**Figure 4.**
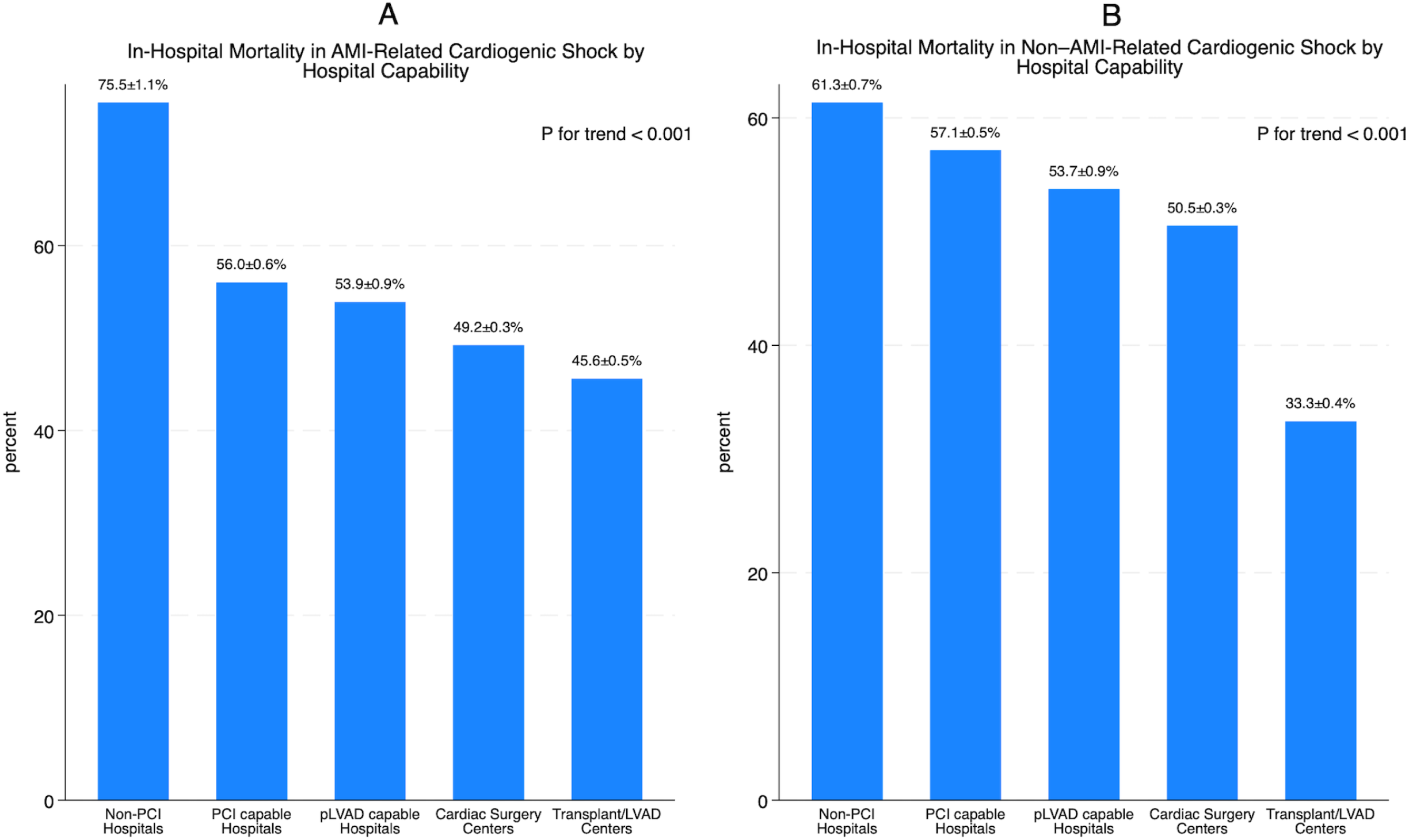
In-hospital Mortality in AMI and Non-AMI CS by Hospital Tier Comparison of unadjusted in-hospital mortality across hospital tiers for (A) AMI-related CS and (B) Non-AMI-related CS. Both cohorts demonstrate a significant stepwise reduction in mortality as hospital capability increases (P for trend < 0.001). Error bars represent 95% confidence intervals.

Within the GSEM for AMI-CS, the latent Acute Severity construct was strongly predicted by AKI, cardiac arrest, and invasive mechanical ventilation, and was the dominant predictor of mortality (Table 2). After adjustment for latent severity, demographics, comorbidities, and hospital characteristics, a graded reduction in mortality persisted across hospital tiers (Figure 5A; Table 2). Additionally, Tier 4 and Tier 5 centers exerted a significant indirect protective effect by reducing Acute Severity among AMI-CS patients.

**Figure 5.**
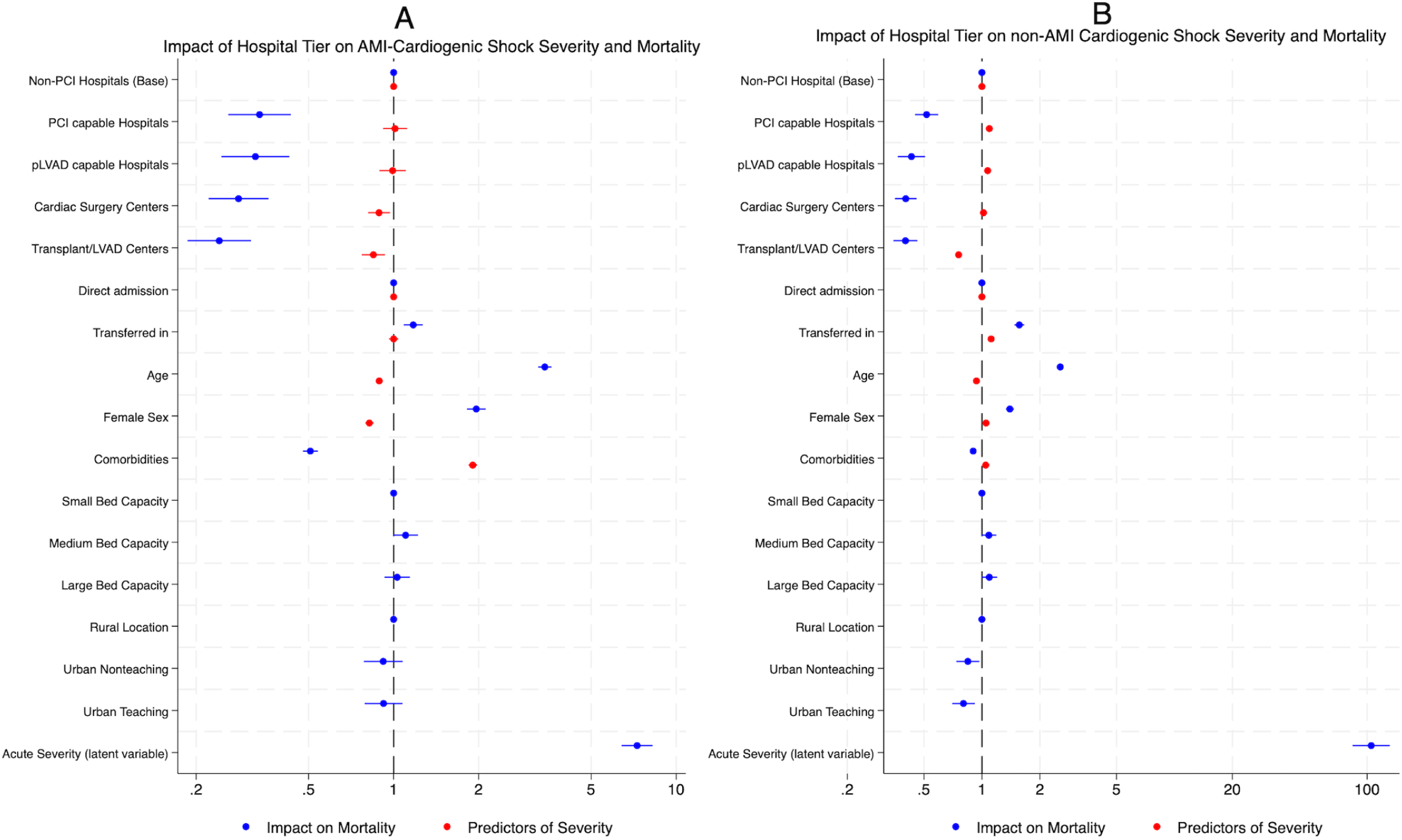
Comparative GSEM Pathways in AMI and Non-AMI Shock (A) AMI-related CS and (B) Non-AMI-related CS. Red dots indicate variables influencing latent severity, and blue dots indicate variables directly associated with mortality. The relative magnitude and direction of associations are shown across both clinical phenotypes. Error bars indicate 95% confidence intervals.

#### Non–AMI-Related Cardiogenic Shock

The remaining 780,455 hospitalizations (66.3% ±0.2%) were for non–AMI-related CS. Unadjusted mortality declined progressively from 61.3% at Tier 1 hospitals to 33.3% at Tier 5 centers (Figure 4B).

In the non–AMI-CS GSEM, Acute Severity was again strongly predicted by AKI, cardiac arrest, and invasive mechanical ventilation, and was an exceptionally strong predictor of mortality (OR 105.1; 95% CI 84.1–131.1) (Table 2). After full adjustment, a stepwise reduction in mortality persisted from Tier 1 through Tier 5 hospitals (Figure 5B; Table 2), with Tier 5 centers also demonstrating indirect mortality benefit through attenuation of Acute Severity.

##### Transfer Status and Hospital Capability

Unadjusted mortality was significantly higher among transferred-in patients compared with direct admissions across all hospital tiers (Figure 6). In the base model, transfer-in status was associated with increased latent severity (OR 1.12; 95% CI 1.10–1.14) and higher in-hospital mortality (OR 1.39; 95% CI 1.33–1.46).

**Figure 6.**
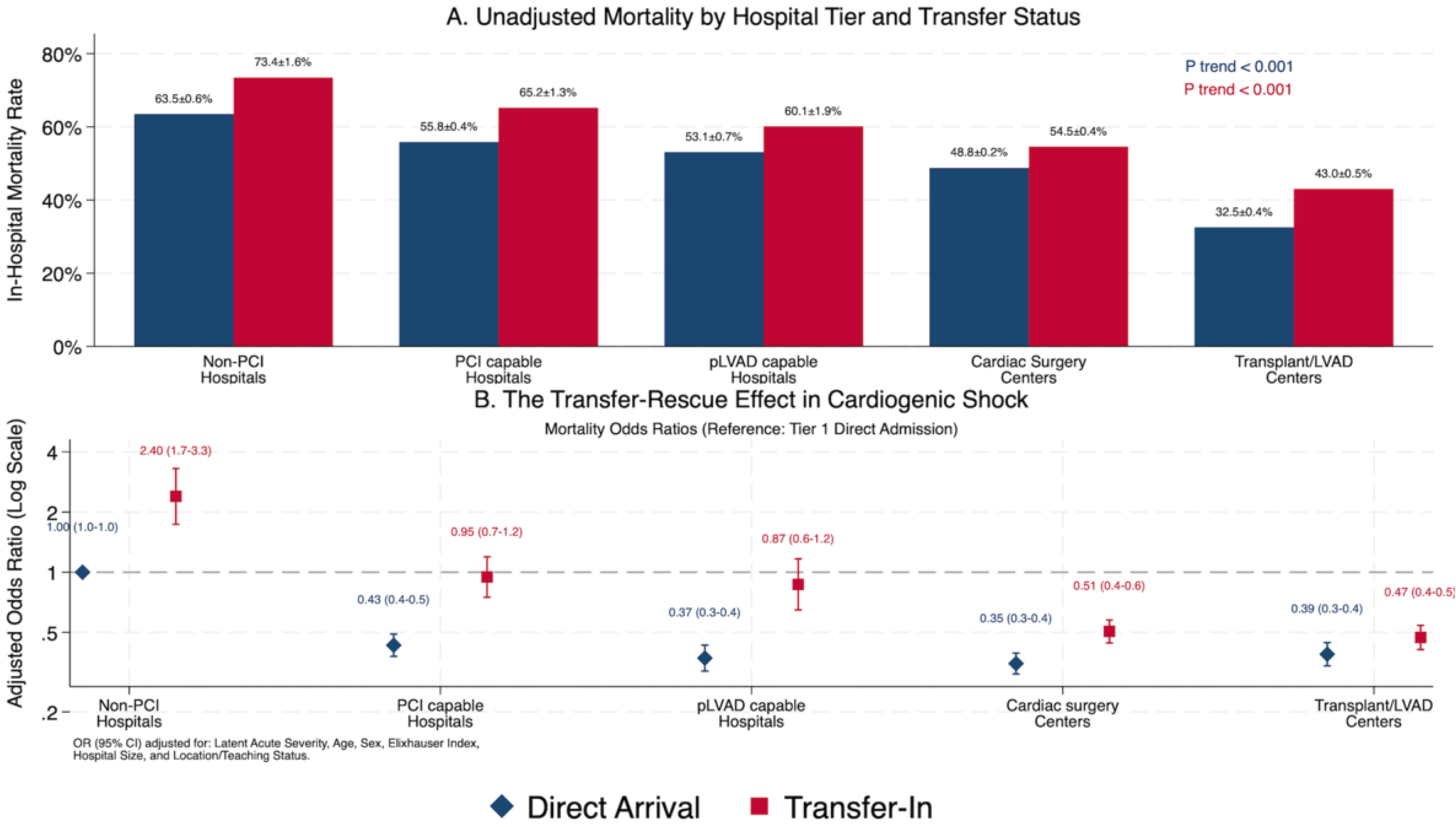
Interaction Between Hospital Tier and Transfer Status Impact of institutional capability on the mortality of transferred patients. (A) Unadjusted mortality rates stratified by hospital tier and patient origin (direct arrival vs. transfer-in). (B) Adjusted odds ratios for mortality illustrating the “Transfer-Rescue Effect”. While inter-hospital transfer is associated with a significant mortality penalty at non-PCI centers (Tier 1), this risk is progressively mitigated at PCI and pLVAD centers (Tiers 2–3) and ultimately reversed at cardiac surgical and transplant hubs (Tiers 4–5). Error bars represent 95% confidence intervals.

After adjustment for Acute Severity and other covariates, a significant interaction emerged between hospital tier and transfer status. While transfer-in status was associated with markedly higher mortality at Tier 1 centers (OR 2.40), this excess risk was significantly attenuated at Tier 4 (Interaction term - OR 0.60 95% CI 0.43-0.84; p=0.003) and Tier 5 centers (OR 0.51 95% CI 0.36-0.70; p<0.001) (Figure 6B), consistent with a “rescue effect” at high-capability quaternary centers.

##### Model Performance and Sensitivity Analyses

All four primary models demonstrated excellent discrimination, with area under the curve (AUC) values ≥ 0.95 (Supplemental Figures S1A-D). To evaluate the robustness of these findings, a series of sensitivity analyses were conducted. First, using standard survey-weighted multivariable logistic regression—adjusting for age, sex, comorbidities, hospital characteristics, and the individual components of the acute severity construct (ALI, AKI, cardiac arrest, and invasive mechanical ventilation)—the inverse association between hospital capability and mortality remained robust (p-for-trend < 0.001).

Consistent with the primary GSEM results, Tier 5 centers were associated with a 61% reduction in the adjusted odds of in-hospital mortality compared with Tier 1 centers (aOR 0.39; 95% CI 0.36–0.42). Furthermore, after re-classifying hospitals using stringent annual volume thresholds (TierAlt: ≥ 10 PCI, ≥ 5 CABG, ≥ 5 pLVAD, and ≥ 2 durable LVAD/transplant), the results remained highly consistent with the primary analysis (Supplemental Figures S2A-B). These secondary models confirm that the observed survival advantage is a stable institutional characteristic of high-capability centers, regardless of the classification methodology employed.

## Discussion

In this large, nationally representative analysis, we identified a robust and stepwise association between hospital cardiac capability and lower mortality in patients with CS. This tier effect persisted after adjustment for demographics, comorbidity burden, and a latent construct of acute physiologic severity—representing the first national demonstration that institutional capability is an independent predictor of outcomes beyond patient acuity alone. While inter-hospital transfer was associated with a crude mortality penalty, this risk was progressively attenuated and ultimately reversed at high-capability centers, suggesting a significant “Transfer-Risk Attenuation Effect” at Tier 4 and Tier 5 institutions.

CS is a complex, heterogeneous syndrome with myriad etiologies and, despite significant advances in cardiovascular medicine, a stagnant mortality rate. CS, secondary to STEMI, is probably the most well studied in randomized controlled trials (RCT). Early revascularization and use of micro-axial pump are the two interventions supported by RCTs and recommended by the current guidelines^13^. However, as noted by prior studies, there is widespread disparities in use of these therapies^6 7^. Recognizing that outcomes are often limited by unequal access to essential infrastructure, specialized multidisciplinary teams, and advanced cardiac resources, the American Heart Association (AHA) has proposed a ‘hub-and-spoke’ regionalization model^9^. This model emphasizes formalized transfer protocols to concentrate high-acuity patients in specialized centers. To empirically evaluate this framework, we developed a hierarchical tier system based on validated administrative coding to characterize institutional cardiac capability. This system serves as a proxy for the presence of a multidisciplinary ‘Shock Team’ and advanced procedural availability, allowing us to quantify how these tiers interact with acute physiologic severity and inter-hospital transfer to determine patient survival^14 15^.

A central challenge in CS research utilizing administrative databases like the NIS is the lack of granular physiological data (e.g., lactate, pH, or cardiac index) to account for clinical instability.

We addressed this by utilizing GSEM to develop a latent “Acute Severity” construct. The validity of this construct as a high-fidelity proxy for shock stage is supported by the heavy statistical loading of its clinical indicators: mechanical ventilation (OR=17.76), cardiac arrest (OR=5.69), and acute kidney injury (OR=2.01). This construct proved to be the most potent predictor of mortality in our model (OR=32.7), successfully “sorting” patients by risk in a way that individual ICD-10 codes cannot (Figure 2 and Table 2).

Our GSEM demonstrated that hospital’s capability influence outcomes by two distinct but complementary pathways: direct effects on mortality and indirect effects mediated through acute physiologic severity (Figure 1). While progressively higher-tier hospitals are associated with lower odds of in-hospital mortality, the attenuation of latent Acute Severity is most pronounced at cardiac surgical and transplant/LVAD centers, suggesting that advanced institutions modify the trajectory of shock rather than simply managing its downstream consequences. Transfer-in status and comorbidity burden increase acute severity, underscoring referral bias toward higher-tier centers, yet this increased severity does not negate the survival advantage conferred by advanced care environments. Notably, the magnitude of association between Acute Severity and mortality far exceeds that of traditional demographic or hospital characteristics, reinforcing that prevention and reversal of multiorgan dysfunction is central to survival in CS.

Our model identified a unique relationship between age and shock dynamics. While increasing age was a direct driver of mortality, younger age was associated with higher latent acute severity (OR=0.92). This likely reflects a ‘salvage paradigm’ in regionalized care, where the younger, critically ill patients are preferentially transferred to higher Tier centers. Female sex was associated with a 61% increase in the adjusted odds of in-hospital mortality (OR=1.61, p<0.001), yet it demonstrated a neutral relationship with the latent Acute Severity construct (OR=1.00).

Consistent with prior studies, this suggests that despite similar clinical instability to men, women experience a significantly higher biological or therapeutic risk^16 17^. Our GSEM identifies comorbidities (Elixhauser comorbidity index) as a powerful upstream driver of acute physiological collapse rather than just an independent marker of mortality. While the direct mortality path for comorbidities appears attenuated, this is due to the dominant mediating role of the latent Acute Severity construct. Essentially, chronic comorbidities increase mortality primarily by predisposing patients to more severe shock trajectories which then serves as the final common pathway for in-hospital mortality. Prior studies have found an association between hospital size and location/teaching status and CS mortality^3^. However, we demonstrated that hospital capability tier independently and more strongly predicts CS mortality. Urban teaching status was associated with lower adjusted mortality, likely reflecting the multidisciplinary infrastructure and 24/7 subspecialty availability inherent to academic medical centers. However, the magnitude of these effects was markedly smaller than the survival benefit conferred by specific cardiac capabilities. Larger bed capacity is associated with a slight increase in the direct odds of mortality compared to small hospitals. This is likely because large hospitals act as regional catchment centers and naturally manage more complex cases that cannot be fully captured even by a latent severity variable.

Consistent with prior single center studies, we found AMI related CS accounted for about one third of all CS visits^8^. Although a tier-dependent survival benefit was observed in both AMI and non-AMI related CS (Figure 4 and 5), important mechanistic differences emerged. In AMI-related CS, advanced hospital tiers primarily conferred a direct survival advantage, consistent with timely revascularization, access to surgical backup, and escalation to temporary mechanical circulatory support. In contrast, in non–AMI CS, Tier 5 centers exerted a disproportionately strong indirect effect by attenuating acute physiologic severity, reflecting the greater dependence of these heterogeneous etiologies on advanced hemodynamic assessment, durable support strategies, and specialized multidisciplinary care. The extreme association between latent severity and mortality in non-AMI shock (OR=105.1) suggests that these patients are clinically ‘labile.’ Unlike AMI-CS, which has a clear culprit lesion for revascularization, non-AMI-CS often represents end-stage system failure that requires the surgical and transplant resources unique to Tier 5 hubs to achieve stabilization.

Although transferred patients experienced higher unadjusted mortality across all hospital tiers—reflecting substantial referral and severity bias—this excess risk was progressively attenuated at higher-capability centers (Figure 6A). After adjustment for latent Acute Severity and other covariates, transfer-in status was associated with markedly increased mortality at non-PCI hospitals, but this risk was neutralized at PCI-capable and pLVAD centers and reversed at cardiac surgical and transplant/LVAD centers (Figure 6B). Notably, the survival association of advanced centers persisted for both direct admissions and transferred patients, indicating that high-capability hospitals not only absorb higher-risk referrals but may actively mitigate the mortality risk associated with interhospital transfer. Together, these findings support regionalized shock systems of care in which early transfer to advanced centers functions as a rescue strategy rather than a marker of futility for the sickest cardiogenic shock patients.

## Limitations

This study has several important limitations. First, the NIS is an administrative database reliant on ICD coding, which is subject to misclassification and cannot capture granular hemodynamic variables such as lactate, pH, or cardiac index. Second, the timing of shock onset relative to admission, transfer initiation, and therapeutic interventions cannot be reliably determined, limiting causal inference regarding transfer timing.

Third, we utilized a ‘presence-of-one’ logic for hospital cardiac capability classification. Institutional capabilities were determined based on procedural codes present within the 20% NIS discharge sample. Although, in high-volume centers performing specialized cardiac procedures, the probability of failing to capture at least one such procedure within a 20% systematic random sample of all hospital discharges is statistically negligible. While misclassification is unlikely in high-volume centers and would bias results toward the null, low-volume centers may be under classified. Fourth, we were unable to directly measure shock team activation, protocolized care pathways, or institutional expertise beyond procedural availability. Fifth, the exclusion of patients transferred out of lower-tier facilities (due to unknown discharge disposition in the NIS) may introduce selection bias. It is possible that the high mortality observed at Tier 1 centers reflects a cohort ‘left behind’ due to perceived futility or instability for transport. Finally, as with all observational analyses, residual confounding may persist despite robust adjustment using latent severity modeling

## Conclusion

In this national analysis of over 1.1 million hospitalizations, increasing hospital cardiac capability demonstrated a robust, stepwise association with lower cardiogenic shock mortality. Beyond direct survival benefits, advanced centers (Tiers 4 and 5) exerted a significant indirect effect by attenuating acute physiologic severity, a phenomenon most pronounced in non-AMI shock. Crucially, while inter-hospital transfer traditionally carries a mortality penalty, this risk was progressively neutralized and ultimately reversed at specialized quaternary hubs, supporting a “Transfer-Rescue Effect”. These findings provide strong empirical evidence for a regionalized “hub-and-spoke” model of care, where early identification and triage to high-capability centers may optimize outcomes for the most clinically complex patients

## Clinical Perspectives

### Medical Knowledge

In a 1.1 million-visit cohort, specialized Tier 5 hubs (Transplant/LVAD centers) were associated with a reversal of the “transfer risk” (OR=2.40 at Tier 1 centers) by attenuating acute physiologic severity (OR=0.74).

### Patient Care

The lower adjusted mortality observed at high-capability quaternary centers may overcome the risks inherent to inter-hospital transport, supporting a strategy of early regionalized triage for cardiogenic shock.

### Translational Outlook

These data provide empirical associations supporting the AHA hub-and-spoke framework, suggesting that formalized regionalization may reduce national cardiogenic shock mortality.

## Data Availability

This data is publicly available from AHRQ.

Central Illustration: The “Transfer Associated Risk” Effect in Regionalized Cardiogenic Shock Care.

Central Illustration summarizes the interaction between hospital capability and survival in 1.1 million cardiogenic shock visits. Initial presentation at Tier 1 centers or inter-hospital transfer is associated with a “Transfer Penalty,” increasing both latent acute severity (OR=1.12) and adjusted mortality (OR=1.39; OR=2.40 at Tier 1). Physiologic instability—represented by the Acute Severity Construct (driven by mechanical ventilation and cardiac arrest)—is the primary determinant of mortality (OR=32.7). High-capability Tier 5 hubs (Transplant/LVAD) provide a potent “Transfer-Risk mitigation” effect through direct mortality reduction (OR=0.35) and the active attenuation of acute severity (OR=0.74), resulting in a final adjusted survival advantage for transferred patients (OR=0.47). These data support a regionalized hub-and-spoke model to reduce mortality from 64.4% to 36.5%.

## Reference

1. Thiele H, Zeymer U, Neumann FJ, et al. Intraaortic Balloon Support for Myocardial Infarction with Cardiogenic Shock. N Engl J Med. 2012;367(14):1287–1296. doi:10.1056/NEJMoa1208410

2. Osman M, Syed M, Patibandla S, et al. Fifteen-Year Trends in Incidence of Cardiogenic Shock Hospitalization and In-Hospital Mortality in the United States. J Am Heart Assoc. 2021;10(15):e021061. doi:10.1161/JAHA.121.021061

3. Vallabhajosyula S, Dunlay SM, Barsness GW, Rihal CS, Holmes DR, Prasad A. Hospital-Level Disparities in the Outcomes of Acute Myocardial Infarction With Cardiogenic Shock. Am J Cardiol. 2019;124(4):491–498. doi:10.1016/j.amjcard.2019.05.038

4. Hochman JS, Sleeper LA, Webb JG, et al. Early Revascularization in Acute Myocardial Infarction Complicated by Cardiogenic Shock. N Engl J Med. 1999;341(9):625–634. doi:10.1056/NEJM199908263410901

5. Møller JE, Engstrøm T, Jensen LO, et al. Microaxial Flow Pump or Standard Care in Infarct-Related Cardiogenic Shock. N Engl J Med. 2024;390(15):1382–1393. doi:10.1056/NEJMoa2312572

6. Nathan AS, Kennedy KF, Reddy KP, et al. Variation in Likelihood of Undergoing Percutaneous Coronary Intervention for ST-Segment–Elevation Myocardial Infarction Among US Hospitals. J Am Heart Assoc Cardiovasc Cerebrovasc Dis. 2025;14(5):e038317. doi:10.1161/JAHA.124.038317

7. Verma A, Chervu NL, Kim JJ, Mallick S, Ziaeian B, Benharash P. Variation in mechanical circulatory support use for acute myocardial infarction cardiogenic shock. Heart Br Card Soc. Published online August 24, 2025:heartjnl-2024-325413. doi:10.1136/heartjnl-2024-325413

8. Berg DD, Bohula EA, van Diepen S, et al. Epidemiology of Shock in Contemporary Cardiac Intensive Care Units. Circ Cardiovasc Qual Outcomes. 2019;12(3):e005618. doi:10.1161/CIRCOUTCOMES.119.005618

9. Contemporary Management of Cardiogenic Shock: A Scientific Statement From the American Heart Association. doi:10.1161/CIR.0000000000000525

10. Garan AR, Kataria R, Li B, et al. Outcomes of Patients Transferred to Tertiary Care Centers for Treatment of Cardiogenic Shock: A Cardiogenic Shock Working Group Analysis. J Card Fail. 2024;30(4):564–575. doi:10.1016/j.cardfail.2023.09.003

11. Quan H, Sundararajan V, Halfon P, et al. Coding algorithms for defining comorbidities in ICD-9-CM and ICD-10 administrative data. Med Care. 2005;43(11):1130–1139. doi:10.1097/01.mlr.0000182534.19832.83

12. Khera R, Krumholz HM. With Great Power Comes Great Responsibility: “Big Data” Research from the National Inpatient Sample. Circ Cardiovasc Qual Outcomes. 2017;10(7):e003846. doi:10.1161/CIRCOUTCOMES.117.003846

13. 2025 ACC/AHA/ACEP/NAEMSP/SCAI Guideline for the Management of Patients With Acute Coronary Syndromes: A Report of the American College of Cardiology/American Heart Association Joint Committee on Clinical Practice Guidelines. doi:10.1161/CIR.0000000000001309

14. Moghaddam N, Van Diepen S, So D, Lawler PR, Fordyce CB. Cardiogenic shock teams and centres: a contemporary review of multidisciplinary care for cardiogenic shock. ESC Heart Fail. 2021;8(2):988–998. doi:10.1002/ehf2.13180

15. Rab T, Ratanapo S, Kern KB, et al. Cardiac Shock Care Centers. J Am Coll Cardiol. 2018;72(16):1972–1980. doi:10.1016/j.jacc.2018.07.074

16. Bansal K, Pawar S, Yamparala A, et al. Sex-Specific Transfer Outcomes in Patients With Non–Acute Myocardial Infarction-Related Cardiogenic Shock. JACC Heart Fail. 2025;13(10):102620. doi:10.1016/j.jchf.2025.102620

17. Ton VK, Kanwar MK, Li B, et al. Impact of Female Sex on Cardiogenic Shock Outcomes: A Cardiogenic Shock Working Group Report. JACC Heart Fail. 2023;11(12):1742–1753. doi:10.1016/j.jchf.2023.09.025

